# Characterizing spatiotemporal patterns of case reporting backfill: a case study of COVID-19 reporting in Michigan, 2020–24

**DOI:** 10.1101/2025.11.25.25340977

**Authors:** Yannan Niu, Andrew F. Brouwer, Emily T. Martin, Joseph R. Coyle, Marisa C. Eisenberg

## Abstract

Backfill is the process of revising case data, often by retrospectively assigning or reassigning newly reported cases to earlier symptom onset dates. Time- and spatial-varying delays in the backfill process may compromise real-time surveillance and forecasting efforts by obscuring the true underlying transmission patterns. Using Michigan COVID-19 case data, we developed a statistical mixture model to describe backfill and geographical and temporal variations. The model combined an exponential process (case reporting delay) and a gamma-distributed process (date reassignment). Parameters were estimated by maximum likelihood with lasso regularization, and the Akaike Information Criterion was used to determine the necessity of the reassignment component for each date. We estimated the exponential reporting speed over time and space and, if appropriate, the transient peak and time of case reassignment. We found that case reporting improved over the pandemic: reporting speed increased over time (with substantial day-to-day variation), and case reassignments were processed faster. We also identified potential regional disparities: rural regions with population densities below 50 people/km^2^ had slower backfill speeds. These findings provide critical insights about the evolution of case reporting and backfill dynamics that can be leveraged for “nowcasting” models to complete real-time surveillance data, ultimately improving outbreak preparedness and response.

## Introduction

Backfill is the process of retrospectively adding or correcting data for earlier dates in epidemiological surveillance systems.^1^ This process of revision is common in infectious disease reporting since it is rarely possible to promptly diagnose and report all cases immediately after symptom onset. *Case reporting delay* is the time between an individual’s symptom onset and that onset date being reported in a case surveillance system. There are multiple factors contributing to this delay, including delays in healthcare seeking, testing, test processing, and disease investigation. Consequently, the case reporting for a particular symptom onset date may remain incomplete for some time, with more recent data typically being more incomplete, i.e., farther away from the *ultimate size* that will eventually be reported for that date. Additionally, in Michigan, cases are usually reported to the surveillance system before disease investigation captures an individual’s onset dates. Therefore, cases are often initially reported with their testing or referral dates (the date when the case is reported into the system) in place of the onset date.^2^ It takes additional time to investigate and identify individuals’ onset dates, after which *case reassignment* to the appropriate onset date can occur. If onset dates cannot be determined, the initial testing or referral dates may remain in the system (Figure 1).

**Figure 1.**
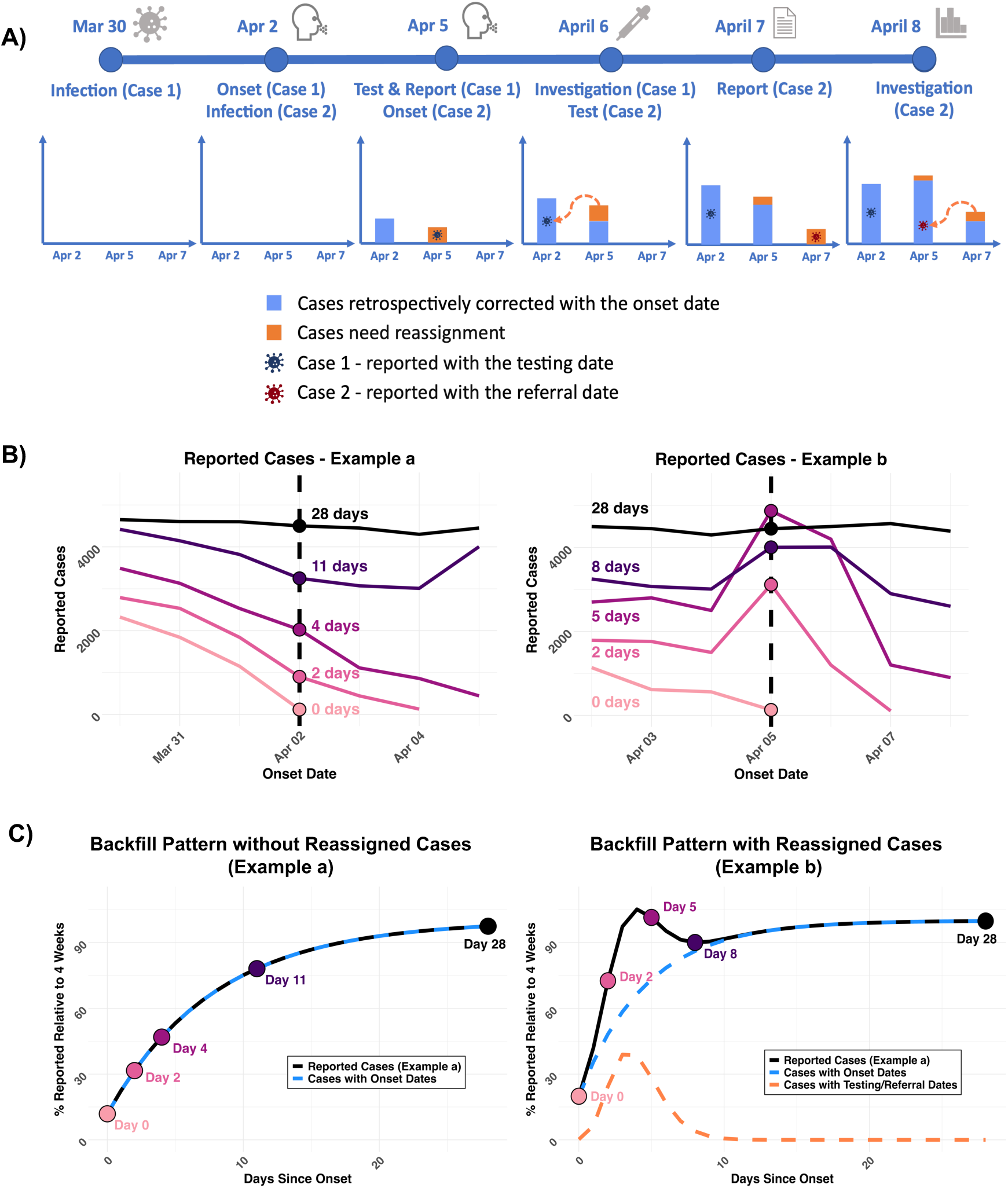
Backfill is the process of retrospectively correcting the number of cases reported for a given symptom onset date. **A)** illustrates the case reporting and investigation timeline. Case 1 was initially reported on April 5 with their testing date and later reassigned to their onset date (April 2) after investigation on April 6. Case 2 was reported on April 7 with a referral date and reassigned to April 5 after case investigation on April 8. By April 8, April 2 only reports delayed-but-corrected cases (with more potentially still to come), while April 5 has both delayed cases and cases requiring reassignment. **B)** shows reported cases by onset date across multiple reporting dates. Each colored line represents the number of cases reported on each onset date after the indicated number of days, progressing from light pink (recent reporting) to black (eventual reporting). Dots mark the reported value for the given reporting date. The vertical dashed lines denote the selected onset dates used in the examples: Example a (left) corresponds to April 2, and Example b (right) corresponds to April 5. **C)** illustrates the backfill patterns corresponding to the examples shown in Panel B, expressed as the percentage of total cases reported by 28 days (4 weeks), plotted over days since the given onset date. Example a (left) shows an exponential accumulation of reports, without the need for reassignment, while Example b (right) includes an additional component (orange line) to account for delayed reassignment of cases. Dots indicate the reported values across reporting dates, labeled by days since onset and colored consistently with Panel B.

The backfill process reflects not only delays in real-time surveillance but also the timeliness and effectiveness of public health responses. Longer delays in backfill may result from delayed disease investigations, particularly during periods of high case volume. Analyzing backfill patterns can provide retrospective insights into case investigation efficiency over time and space. Additionally, many studies have recognized that backfill delays considerably compromise forecasting model performance and intervention evaluation.^1, 3-6^ These studies have proposed several methods to mitigate the impact of backfill, including using up-to-date data by tracking all revisions from various data sources, correcting recent data with nowcasting models, or accounting for backfill dynamics within their models to enhance prediction.^1, 4, 7^ While these techniques can help to improve model performance, they are usually based on assumptions of a consistent backfill process.^6^ However, backfill can vary across time and regions because of the differences in public health infrastructure and reporting capacity.^8^ Not accounting for these potential patterns would reduce the accuracy and utility of nowcasting and other correction techniques.

COVID-19 case data provide an opportunity to comprehensively explore backfill patterns because of their high quality and extensive volume during the pandemic. This study aims to characterize the backfill process and its dynamics and spatiotemporal patterns in COVID-19 case data in Michigan, 2020–22. We developed a statistical model to describe backfill patterns as a function of case reporting and reassignment, and we estimated the model parameters, characterizing backfill dynamics across spatial regions and over time. Our findings will provide valuable insights into backfill dynamics, uncovering how case delay and reassignment influence real-time surveillance over time and space.

## Methods

### (1) Data source

COVID-19 case data were obtained from the Michigan Disease Surveillance System. This analysis included confirmed cases by onset date from May 1^st^, 2020, to September 24^th^, 2024, from 83 counties and the city of Detroit. Cases without specified geographical information were categorized as “Unknown County”. State-level data aggregated cases from 85 areas (83 counties, Detroit, and Unknown). Regional-level data aggregated cases into 8 emergency preparedness regions (1, 2N, 2S, 3, 5, 6, 7, 8), grouped based on the state emergency response procedures and protocols, and excluded cases from “Unknown County”.^9^ Regional population density was calculated with county-level population statistics, sourced from the Michigan Department of Health and Human Services.^10^ Because this study used aggregated data for public health surveillance, this study was not regulated as human subjects research (University of Michigan Health Sciences and Behavioral Sciences Institutional Review Board HUM00181319).

The pandemic period was subdivided into five periods: the Original, Alpha, Delta, Omicron waves, and the post-pandemic period. The dates of the first four waves were determined by the proportion of variants at the state level, with cut-offs of May 1^st^, 2020 (the first date in the dataset), March 7^th^, 2021, June 28^th^, 2021, and December 13^th^, 2021, respectively.^11^ The start of the post-pandemic period was determined by the end of the federal COVID-19 emergency declaration on May 11^th^, 2023.^12^

### (2) Ultimate size and normalization

The number of cases reported for a specific onset date usually stabilizes at its ultimate size after several weeks of reporting and possible reassignment. In this study, the ultimate size for a given onset date was defined as the reported case count by 28 days after onset. A four-week period was sufficient to capture most short-term corrections, reflecting the natural backfill dynamics.^7^ Simultaneously, it excluded irregular corrections that happened months or years later due to changes in case definitions or policy changes. Case counts, from 0 to 28 days after each onset date, were then normalized by their ultimate sizes, standardizing backfill as a fraction for direct comparisons across dates, regardless of changes in case volume.

### (3) Empirical analysis

To characterize the spatiotemporal variations in backfill patterns, we visualized the distributions of normalized case counts for each onset date across various dimensions, including pandemic waves, geographical levels, release frequencies, and ultimate sizes. The five waves and geographical levels were specified above in the Data Source sections. Release frequencies were cut as 5-7 times, 3-4 times, 2 times, and 1 time per week. Ultimate sizes were classified into <100, 100-1,000, 1,000-5,000, 5,000-10,000, and >10,000.

### (4) Model

We developed two backfill models to capture observed patterns: a) a model accounting only for delayed reporting and b) a model with both delayed reporting and case reassignment.

#### a) Reporting-only model

Delayed reported cases initially increase rapidly and gradually stabilize at their ultimate sizes (Figure 1C: Example a). To capture this pattern, we developed a reporting-only model using an exponential process (Equation 1), where *t* represents the number of days since onset (with *t*=0 being the onset date) and *ε* denotes the random error. The reporting-only model assumed that all delayed reported cases were added at a constant exponential reporting speed *λ* without reassignment, i.e., cases are only reported once their symptom onset dates are available. A larger value of the reporting speed might indicate a more efficient case-reporting process, which captures all cases in a shorter period. This process, though simple, adequately described the sharp initial rise and subsequent slowing down and stabilization observed for many dates.

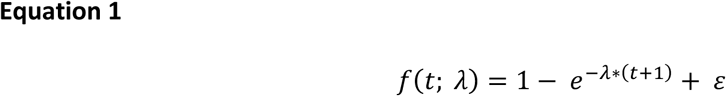

#### b) Reporting–reassignment model

The reporting-only model does not account for case date reassignment and thus cannot fit the observed backfill patterns for many dates in our data (e.g., Figure 1C: Example b). We developed a reporting-reassignment model by adding a gamma-distributed process of case reassignment to the reporting-only model (Equation 2). In this model, *g*(*t*; *κ*, *θ*) is the probability density function of the Gamma distribution: 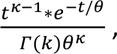 where 𝛤(*k*) is the Gamma function. As in the reporting-only model, the parameter *λ* represents the reporting speed. The reporting-reassignment component includes three additional parameters: *β*, *k*, and *θ*, representing the relative weight, shape, and scale of the gamma-distributed reassignment process, respectively. We derived two summary measures from the above three parameters to characterize the reassignment pattern - transient peak and peak time. The transient peak of the reassignment distribution denotes the maximum proportion of reassigned cases relative to the ultimate reported case counts. A lower peak value reflects a relatively smaller maximum volume of reassigned cases. The peak time reflects the timing of the transient peak after which removal becomes the dominant trend.

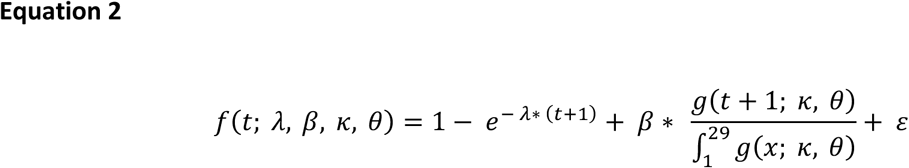

### (5) Model fitness and parameter estimation

We estimated parameters with maximum likelihood estimation by minimizing the negative log-likelihood (NLL) (Equation 3) with both models. To ensure global minimization, we explored a wide range of initial values for *κ* (1 to 100) and *θ* (1 to 20) to avoid local minimization due to improper initial values. However, reporting and reassignment did not occur uniformly across all dates, and the reassignment component was not necessary to describe the observed backfill for every date. Therefore, we used the Akaike Information Criterion (AIC), *AIC* = 2*n* − 2 *log L*, where n is the number of parameters, incorporating both model complexity and likelihood, to determine whether adding the reassignment component provided a better fit to the observed backfill pattern on each date.

Although these models reasonably described most patterns in the data, the reporting-reassignment model overfitted the case data on some dates, resulting in overly large *β* and corresponding extremely small *λ*, lacking practical interpretability. To mitigate overfitting and improve interpretability, we added a lasso penalty term on *β* (*μ* ∗ |*β*|) into the NLL, where *μ* was the regularization coefficient, to constrain large values of *β*. We explored choices of the coefficient *μ* from 0 to 0.5 (Supplementary Table S1). A larger *μ* corrected extremely low reporting speed *λ* caused by overfitting, while making the speeds for some days appear faster than they actually are. For the main results, we set the regularization coefficient to 0.3 since this value effectively fixed most abnormal parameters without excessively distorting them. The penalty term was not included when calculating the AICs.

Model parameters were estimated for each onset date to summarize the backfill process, including the reporting speed, and, if relevant, the transient peak and peak time of the reassignment component. However, changes in the frequency of case data release substantially impacted the data availability for parameter estimation. During the pandemic, release frequency shifted gradually from daily to weekly (Supplementary Figure S1), resulting in fewer than 10 data points per onset date in 2022. Beyond April 2022, the reduced frequency, fewer than 3 times per week, limited reliable parameter estimation. As a result, model parameters were estimated for dates between May 1, 2020, and April 30, 2022, only.

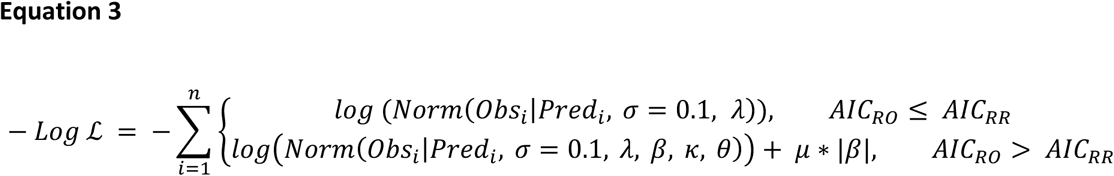

## Results

### (1) Empirical distributions of backfill

The distributions of COVID-19 case reporting data across the five waves had similar patterns, but with variations (Figure 2A-E). In all waves, normalized cases across all dates in that wave showed a rapid increase in the first week and gradually stabilized around 1. However, their specific slopes and quantiles were not consistent. Curves became steeper over time, especially after the Alpha wave. The steeper curves suggested an accelerated backfill process in later waves, requiring less time to reach stabilization. The widths of the quantiles generally became wider, especially during the Omicron wave and post-pandemic, indicating less consistent backfill patterns. Simultaneously, the impact of reassigned cases became more obvious after the Delta wave, with normalized cases frequently exceeding their ultimate sizes (i.e., normalized case volumes over 1) at points over the 4-week period. In addition, backfill patterns varied across different ultimate sizes (Supplementary Figures S2 and S3). The general reporting speed did not show significant differences across ultimate sizes, but the slopes were gentler for dates with smaller ultimate sizes between 100 and 1,000, where most dates in 2020 were concentrated, suggesting a slower reporting speed (Supplementary Figures S3). While ultimate sizes were related to time periods, the 75%, 85%, and 95% quantiles still became tighter as the ultimate size increased, especially when they exceeded 10,000. This pattern suggested large case counts might mitigate the impact of stochasticity in reporting and increase consistency across onset dates. Data release frequencies reduced from daily to weekly throughout the pandemic (Supplementary Figure S1), with significant decreases as described in the Methods. However, backfill distributions were largely consistent within a period, regardless of variations in reporting frequencies (Supplementary Figure S4).

**Figure 2.**
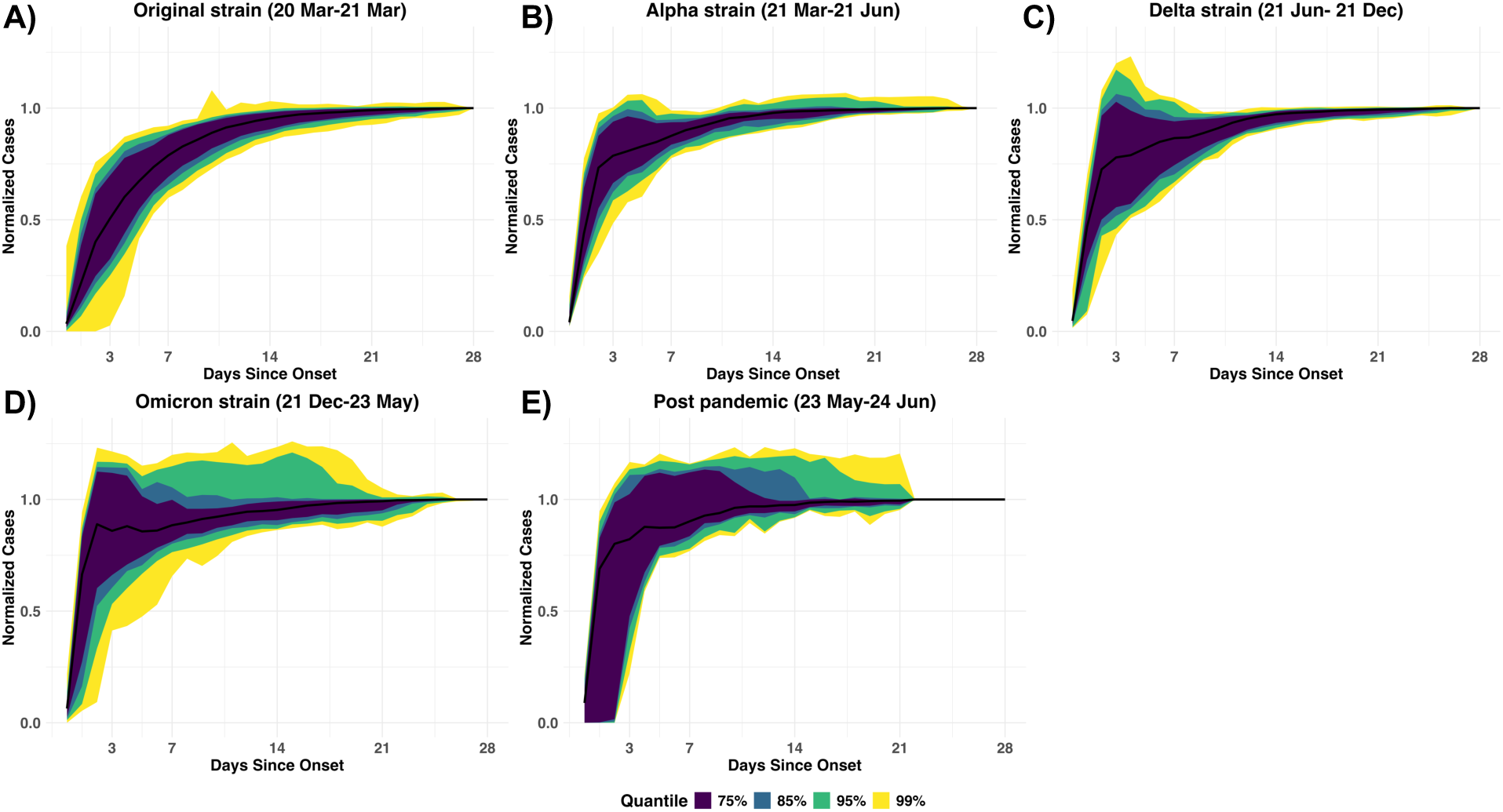
Distribution of COVID-19 case backfill patterns across waves. **A) – E)** shows the distribution of COVID-19 case backfill in the Original, Alpha, Delta, Omicron, and post-pandemic waves, respectively. Four colors represent 75%, 85%, 95%, and 99% quantiles.

At the regional level, backfill patterns exhibited similar trends to the state-level ones overall but varied across eight emergency preparedness regions in the state. In general, backfill curves became steeper over time, and reassigned cases were more apparent in later waves, similarly to state-level patterns (Supplementary Figure S5). However, regional backfill patterns typically showed wider quantiles than state-level, especially in the post-pandemic wave when case counts were limited. Additionally, backfill distributions in the most rural regions, Regions 7 and 8 (Northern Michigan and the Upper Peninsula), showed notably gentler slopes and wider quantiles, indicating slower reporting speeds and less consistent data. This variability might be due to differences in case counts, case processing capacities, or reporting efficiency between the rural and urban areas.

### (2) Parameter estimation

At the state level, the need for the reporting-reassignment model compared to the reporting-only model changed over time. The reporting-only model was selected more frequently during the Original wave (May 2020-March 2021) and became less prevalent later (Figure 3A). In the Original wave, the reporting-only model was selected by more than 75% of dates in each month. In contrast, it was selected for only about 50% of dates by March 2021 and further dropped to nearly 25% by February 2022, though it briefly rose to 25% again in January 2022. Therefore, the reassignment component improved model fitness substantially in later waves. At the regional level, the reporting-reassignment model took a larger percentage compared to the state level (Supplementary Figure S6). In particular, at the aggregated state-level, backfill patterns were more likely to be impacted by urban regions with larger populations, like Regions 2N and 2S (Southeast Michigan, including Detroit City). However, a relatively small number of reassigned cases can significantly impact model selection for regions with a limited population but only have a negligible effect at the state level due to its larger population. As a result, model selection was distributed differently at the regional level from the state level—this also illustrates the importance of exploring more local or regional backfill patterns to understand trends.

**Figure 3.**
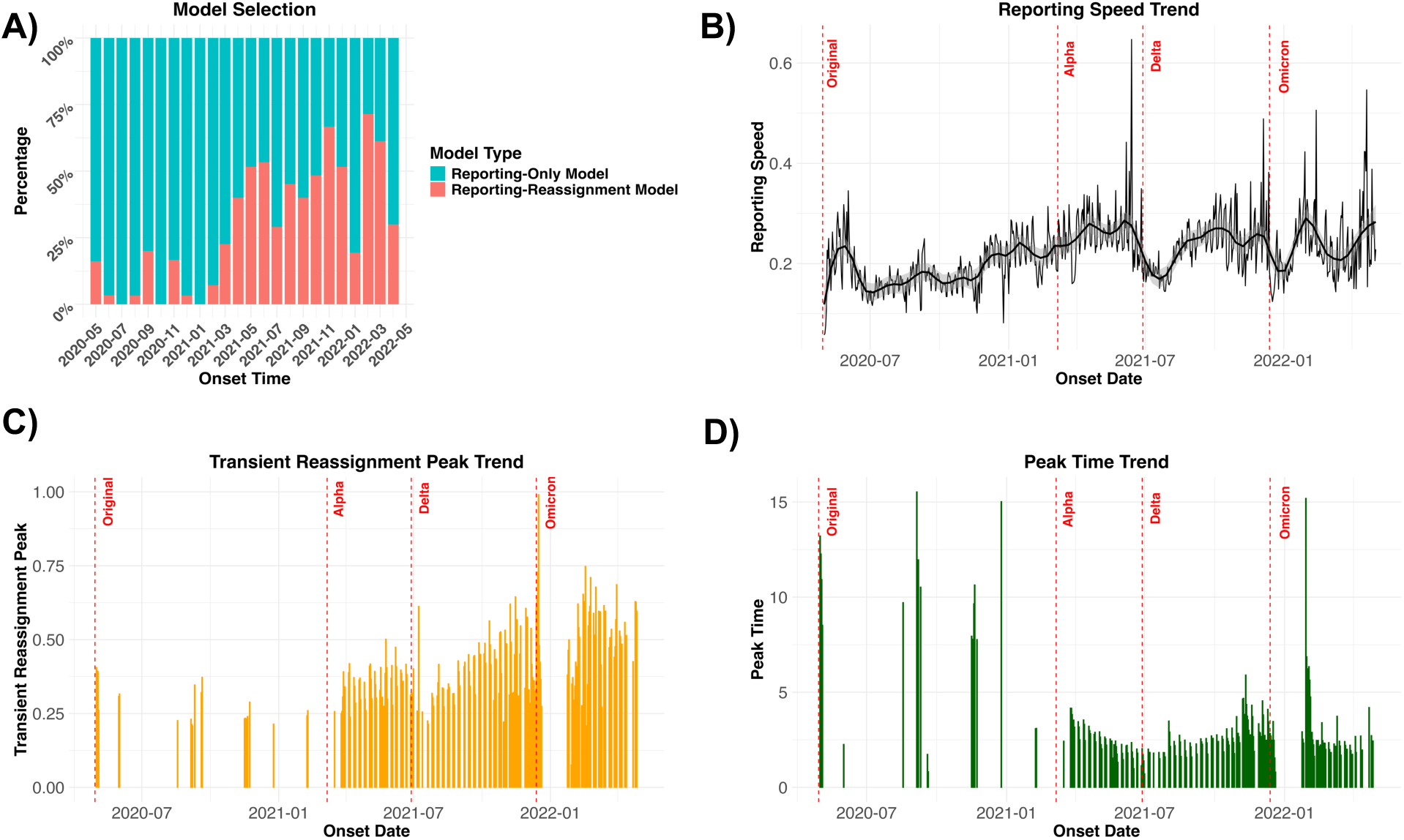
State-level model selection and parameter trends over time. **A)** Monthly percentages of the reporting-only and reporting-reassignment models, with two colors representing the different models. **B)** Reporting speed trends, with a line for the daily parameter and a smoothed line for the overall trend. **C) – D)** Trends of transient reassignment peak and peak time, with blanks representing dates selecting the reporting-only model. Four vertical red dashed lines mark the start of a new wave in panels (B-D).

The magnitude of the reporting speed generally increased over the two years (from ∼0.15 in 2020 to 0.25-0.30 during the Alpha wave), with several intermittent decreases (Figure 3B). While some individual rates exceeded 0.30 afterward, the overall trend, as indicated by the smoothed curve, remained around 0.30 in the Delta wave and portions of the Omicron wave. Translating these values into interpretable terms, it would take about 15 days to capture 90% of cases for each onset date with a reporting speed of 0.15, but it would take less than 10 days for a rate of 0.25. In addition to this overall increasing trend, there were three short-term decreases in the exponential rate occurring in June-July 2021, December 2021, and March-April 2022, each dropping to nearly or even below 0.2.

The process of reassigning dates was measured with the transient reassignment peak and peak time (Figure 3C-D). Transient peaks were low during the Original wave, with the maximum volume of reassigned cases taking up about 25% of the ultimate size. Peaks increased later, exceeding 50% of the ultimate size (Figure 3C). It usually took more than 10 days to reach peak reassignment during the Original wave, while the peak time decreased to approximately 3 days by the Omicron wave (Figure 3D). These trends together suggested that reassignment patterns for cases shifted across waves. In the Original wave, case reassignment occurred less often and more slowly, while in the Alpha and Delta waves, reassigned cases were more common and removed more quickly, within the first week after onset, contributing to a higher peak and shorter peak time. During the Omicron wave, although these cases were still processed rapidly, they contributed larger peaks compared to the Alpha and Delta waves, likely due to the overwhelming case counts at that time.

The regional reporting speeds had similar temporal trends to the state-level ones but varied in magnitude across regions (Figure 4). Most regions showed reporting speeds similar to the overall state, but Regions 3, 7, and 8 (Mid-East and Northern Michigan and the Upper Peninsula) showed relatively low rates, especially during the Original and Alpha waves. Specifically, the reporting speed in Region 7 (Northern Lower Peninsula) was approximately 0.05 lower than other regions, which translates to approximately an additional eight days to capture 90% of cases. These lower rates suggested a less efficient backfill process. Disparities in reporting speeds aligned with regional average population densities (Figure 4A). Population densities in Regions 3, 7, and 8 were lower than 50 people/km^2^, with 48, 20, and 7 people/km^2^, respectively. In contrast, Region 2S, which includes the city of Detroit, had the highest population density of 470 people/km^2^. Nevertheless, this alignment between population density and reporting speed disappeared in the Omicron wave. Regional parameters of reassignment – transient peak and peak time – followed their general temporal trends at the state-level but with larger values, indicating the larger impact from case reassignment within regions (Supplementary Figure S7-S8). These parameters from rural Regions 7 and 8 had the highest values, indicating a larger relative volume of reassigned cases and a longer reassignment process, contributing to a less efficient backfill pattern.

**Figure 4.**
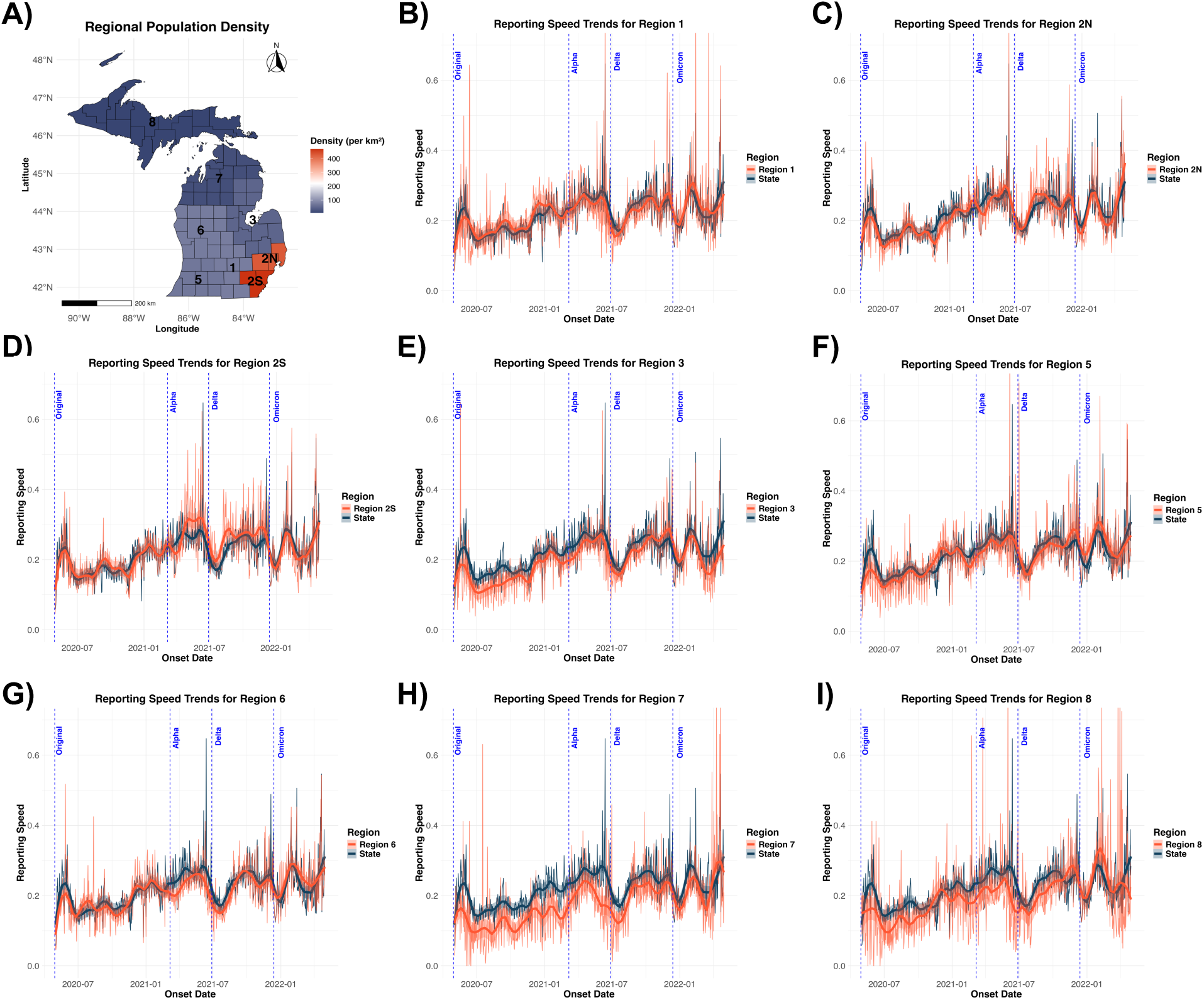
Regional population density and reporting rate trends. **A)** A map of Michigan showing regional population density, with red for higher density and dark blue for lower density. **(B-I)** Exponential trends for eight emergency preparedness regions. The red line represents the regional trend, while the dark blue line represents the state-level trend, serving as a reference. Four vertical blue dashed lines mark the start of a new wave. The reporting speed was lowest overall for Regions 3, 7, and 8 (E, H, I), the regions with the lowest population densities.

## Discussion

Using a statistical mixture model combining an exponential reporting process and a gamma-distributed case-reassignment process, we effectively characterized the backfill process and distinguished delayed case reporting from case reassignment from the initially reported testing or referral dates to symptom onset dates. The backfill process for COVID-19 cases in Michigan improved throughout the pandemic, showing higher efficiency in both delayed reporting and case reassignment. Nevertheless, while reporting efficiency improved overall, there were fluctuations over time, which may be difficult to predict and account for in nowcasting models. We also highlighted spatial heterogeneity, with slower reporting in less population-dense regions compared to more population-dense regions.

Over the first two waves of the pandemic (Original and Alpha), the reporting speeds and transient reassignment peaks gradually increased, and the peak time of reassignment decreased, suggesting improvements to backfill processes. During this period, there was a substantial increase in epidemiological case reporting infrastructure and laboratory testing capacity, which improved the efficiency of case investigation and reporting.^13-15^ Following the initial surge, the CDC issued guidance for submitting standardized electronic laboratory reporting files, allowing non-traditional laboratories to report directly to state health departments.^16^ In parallel, health departments expanded their workforce to meet the rapidly growing demands of the pandemic response.^17^ Simultaneously, the growing understanding of disease transmission facilitated case investigation.^18^ However, the reporting speed did not continue to increase indefinitely, eventually fluctuating around 0.25. This stabilization of the reporting highlights the fact that some degree of delayed reporting will always be unavoidable, even with effective testing techniques and reporting infrastructure.

The overall trends in improvement of backfill efficiency were not monotonic. Several notable downward trends occurred over the two years in the reporting speed, especially in the Delta and Omicron waves. It is possible that the overwhelming volume of cases strained the reporting infrastructure during these periods.^19^ In Michigan, the Omicron wave followed the Delta wave closely, occurring back-to-back without a meaningful pause, which increased the burden on the surveillance system.^20^ Despite a higher case volume than the Delta wave, the Omicron wave showed a shorter dip in the reporting speed. However, the higher reporting speeds in the Omicron wave may not purely indicate better performance in onset date entry. Instead, the huge volume of cases likely exceeded the capacity of timely processing and investigations. As a result, many cases may have remained with their referral or testing dates without undergoing reassignment. This interpretation is supported by the low proportion of days in January 2022 that were better explained by the reporting-reassignment model. The lack of reassignment likely inflated estimates of the reporting parameter, which should not be interpreted as a true reporting improvement (if the goal is representing cases by their onset date). Additionally, reduced data release frequency before the Delta and Omicron waves may have prolonged the system’s adaptation to surging cases, further contributing to observed dips. These patterns suggest that no single factor can fully explain the observed temporal variations; instead, the coincidence of surging case volumes and reduced release frequency may have jointly contributed to challenges in timely case reporting.

We found that regional backfill patterns were broadly similar, with reduced efficiency observed in some rural areas having lower population density. However, the relationship between efficiency and population density was not linear. In our study, substantially lower reporting speeds were observed only in the three lowest population density regions (all have quite low densities below 50 people/km^2^). Michigan’s centralized case investigation workforce during the pandemic likely supported consistent reporting patterns across most regions. Nevertheless, some rural settings may still have faced case reporting challenges. In these areas, healthcare is less accessible, delaying the case diagnosis and reporting.^21^ Additionally, demographic factors impacting investigation priorities, such as age-related targeting strategies informed by urban region data, and disparities in reporting infrastructure may still have led to variations in these rural regions.^22, 23^ Taken together, the disparity may suggest that case reporting challenges present an apparent influence on backfill efficiency when population density falls below a specific threshold.

Our results revealed important spatiotemporal variations in backfill patterns, highlighting that nowcasting models may need to account for variation in time and space. These variations could reduce the accuracy of fixed nowcasting models commonly used in current studies, especially when backfill patterns change dramatically from the initial period. Therefore, a time-varying nowcasting model would be necessary to adapt to shifting backfill characteristics throughout an outbreak. Additionally, the regional disparities also suggest a more granular approach to get more accurate estimates may be needed, especially for rural areas with lower reporting efficiency. Future work could develop a nowcasting model that is both time-varying and region-specific, leveraging backfill dynamics to correct incomplete reported case data. This approach will be instrumental in improving real-time surveillance and supporting outbreak management.

Three aspects require caution when interpreting results from reporting models. First, selecting the reporting-only model does not imply a complete absence of reassigned cases. Instead, it reflects that they are too limited to justify a more complex model within a large population. For example, small numbers of reassigned cases can justify the reporting-reassignment model in rural regions during the Original wave. However, they are insufficient to shift model preference at the state level for the same date, resulting in a higher proportion of the reporting-only model. Secondly, interpreting parameters of individual dates can be misleading because imperfect empirical data on certain days may produce extreme parameter values without meaningful interpretability. For instance, abnormally high reporting speeds exceeding 5 can result from a large number of cases reported on day 0. Nevertheless, this pattern happens rarely in practice, affecting less than 0.5% of all days. To avoid misunderstanding, it is crucial to focus on the larger trends across a period of days instead of overemphasizing an individual daily estimate. Thirdly, case counts reported on the same day as symptom onset (Day 0) are usually less reliable than subsequent days. Though the sensitivity analysis (see Supplementary material) suggests that including Day 0 data can yield more meaningful parameter estimates than excluding it entirely, relying only on Day 0 counts for nowcasting often leads to an underestimation of the ultimate size. Therefore, we should avoid using Day 0 data by itself for real-time estimation when correcting reporting delays.

In conclusion, both case reporting and case reassignment are critical processes to account for in the backfill process, and we developed a statistical mixture model to mechanistically account for both processes. The model effectively captured both delayed case reporting and retrospective corrections on cases initially reported with testing or referral dates. By evaluating the COVID-19 case backfill performance in Michigan, we found overall improvement in reporting efficiency over time but intermittent variations, particularly at the beginning of the Delta and Omicron waves. We also found that rurality impacted reporting speed, with regions with lower population density reporting more slowly. These findings provide valuable insights into backfill dynamics, uncovering how delayed and reassigned cases influence real-time surveillance. The model can also be adapted to other reportable infectious diseases, with appropriate modifications based on their specific reporting mechanisms. By leveraging these insights, “nowcasting” models can predict accurate case estimates with early reported data, addressing reporting delays and corrections, and ultimately improve outbreak preparedness and response.

## Data Availability

All data produced in the present study are available upon reasonable request to the authors and IRB approval and/or a data use agreement from the Michigan Department of Health and Human Services may be required.

## Acknowledgements

This publication was made possible by the Insight Net cooperative agreements NU38FT000002 from the CDC’s Center for Forecasting and Outbreak Analytics (CDC-RFA-FT-23-0069). Its contents are solely the responsibility of the authors and do not necessarily represent the official views of the Centers for Disease Control and Prevention. We thank Julie Gilbert for her assistance with data processing.

## Notes

### Competing Interest Statement

The authors have declared no competing interest.

### Author Declarations

The Health Sciences and Behavioral Sciences Institutional Review Board of the University of Michigan waived ethical approval for this work.

