## Supplementary Materials for "Characterizing spatiotemporal patterns of case reporting backfill: a case study of COVID-19 reporting in Michigan, 2020–24"

Yannan Niu[1], Andrew F. Brouwer[1]*, Emily T. Martin[1], Joseph R. Coyle[2], Marisa C. Eisenberg[1]*

[1] Department of Epidemiology, School of Public Health, University of Michigan
[2] Michigan Department of Health and Human Services
* Contributed equally to this work

**Supplementary Materials:**

1. **Supplementary Table S1 -** Summarization of Results Across Regularization Coefficients – Page 2
2. **Supplementary Figure S1** - COVID-19 case data release frequency trend – Page 3
3. **Supplementary Figure S2** - COVID-19 case backfill ultimate size – Page 3
4. **Supplementary Figure S3** - Distribution of COVID-19 case backfill across ultimate sizes – Page 4
5. **Supplementary Figure S4** - Distribution of COVID-19 case backfill under different reporting frequencies – Page 5
6. **Supplementary Figure S5** - Regional distribution of COVID-19 case backfill across waves – Page 6
7. **Supplementary Figure S6** - Model selection across regions – Page 7
8. **Supplementary Figure S7** - Transient reassignment peak trends across regions – Page 8
9. **Supplementary Figure S8** - Peak time trends across regions – Page 9
10. **Supplementary Analysis** - Sensitivity analysis methods and results – Page 10
11. **Supplementary Figure S9** - Model selection and exponential rate from sensitivity analysis – Page 11
12. **Reference** – Page 12

**Supplementary Table S1 Summarization of Results Across Regularization Coefficients**

| $\mu$ | No. Basic[1] | No. Exp-Gamma[2] | No. $\lambda < 0.1$[3] | No. $\lambda > 0.4$[4] |
|---|---|---|---|---|
| 0.00 | 455 | 275 | 41 | 9 |
| 0.05 | 467 | 263 | 30 | 9 |
| 0.10 | 479 | 251 | 25 | 9 |
| 0.15 | 496 | 234 | 16 | 9 |
| 0.20 | 503 | 227 | 12 | 10 |
| 0.25 | 509 | 221 | 10 | 10 |
| **0.30** | **518** | **212** | **4** | **10** |
| 0.35 | 530 | 200 | 4 | 12 |
| 0.40 | 530 | 200 | 4 | 13 |
| 0.45 | 535 | 195 | 4 | 13 |
| 0.50 | 541 | 189 | 3 | 15 |

[1] The number of dates selected the basic model out of 730 dates over two years.

[2] The number of dates selected the exponential-gamma model out of 730 dates over two years.

[3] The number of dates with the exponential rate below 0.1, indicating extremely slow reporting speed.

[4] The number of dates with the exponential rate above 0.4, indicating extremely fast reporting speed.

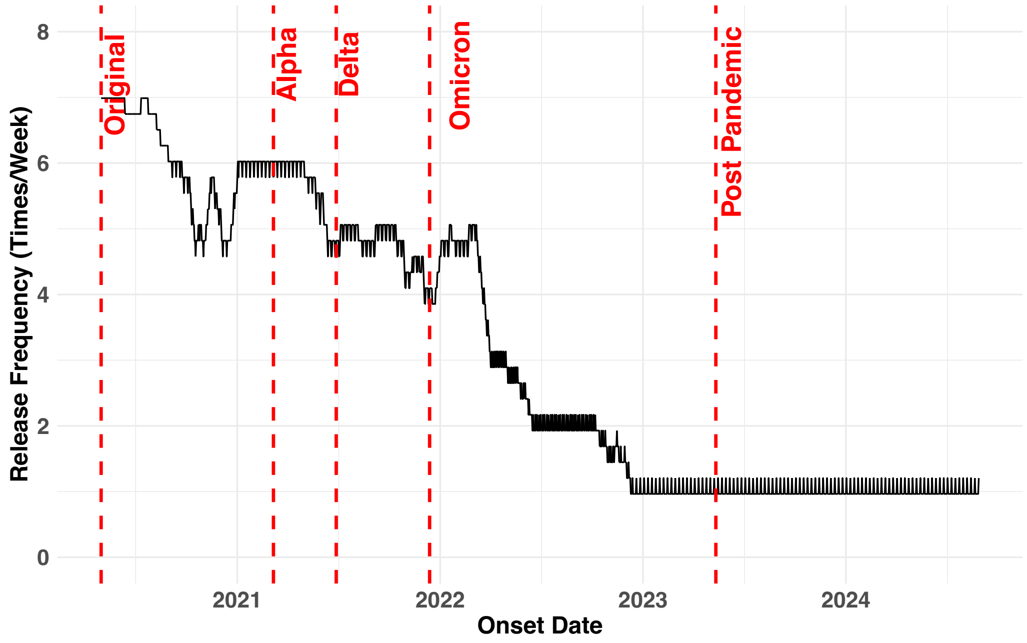


**Supplementary Figure S1** COVID-19 case data release frequency trend. Displaying the trend of the frequency of data release over time, with five vertical red dashed lines marking the start of a new wave

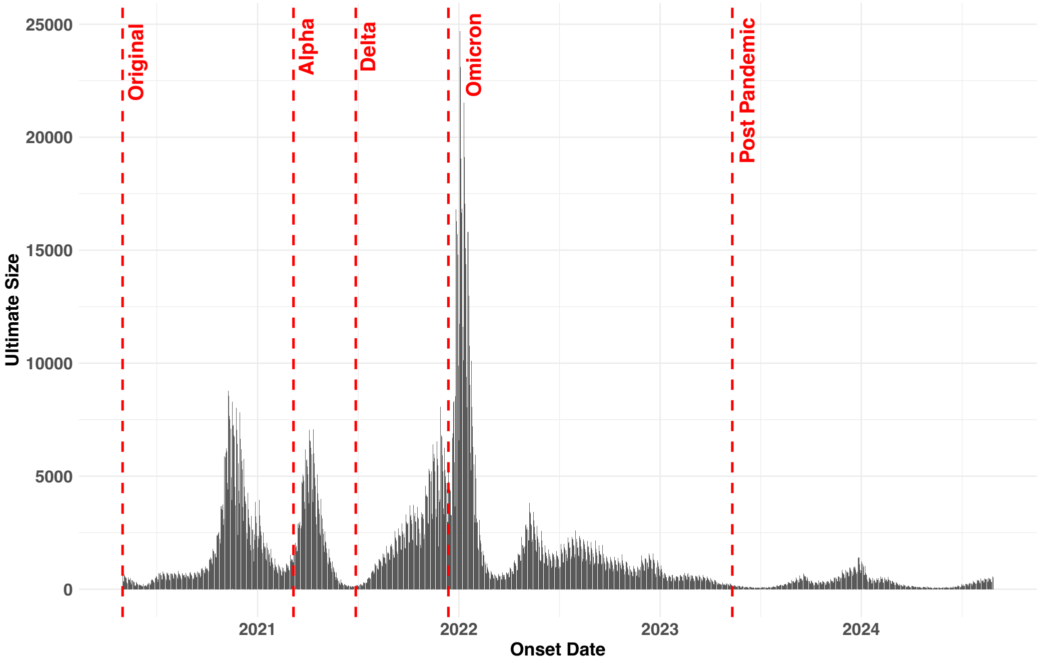


**Supplementary Figure S2** COVID-19 case backfill ultimate size. Displaying the ultimate sizes of COVID-19 cases reported 28 days after onset for each onset date. Five vertical red dashed lines mark the start of a new wave.

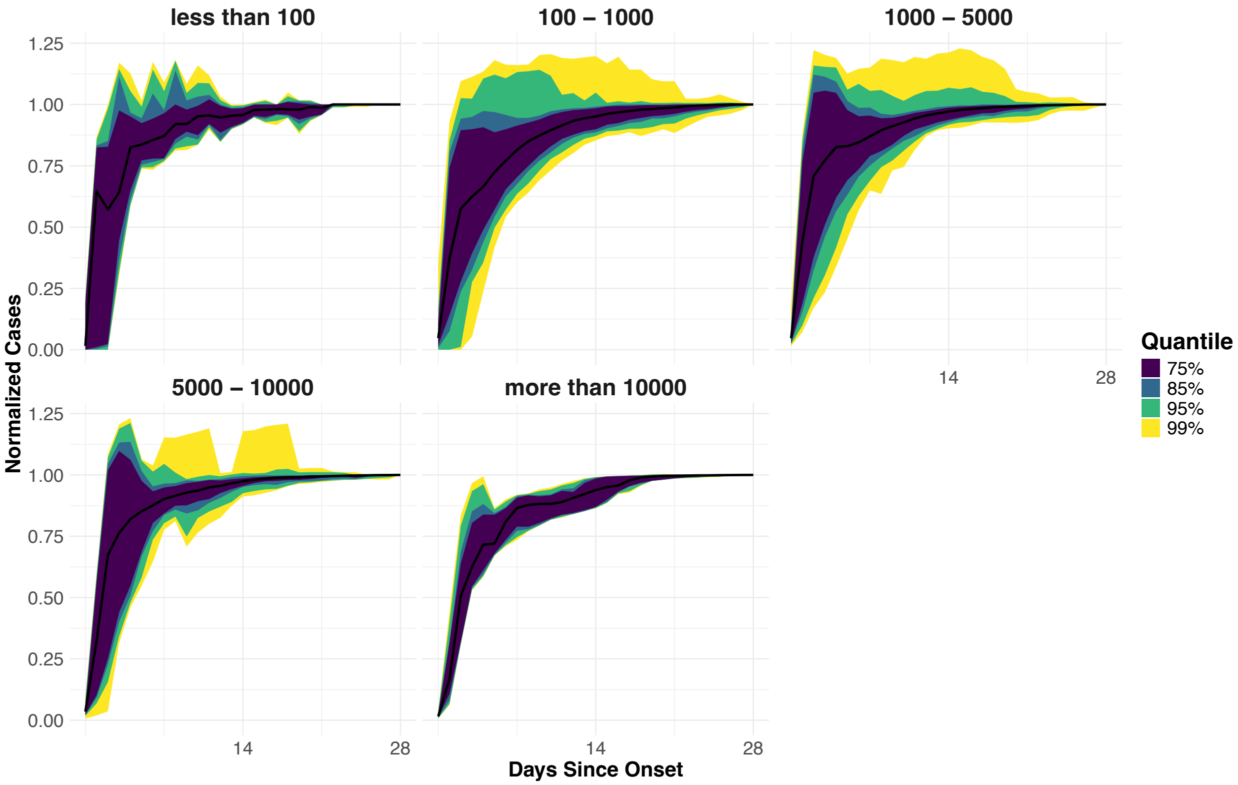


**Supplementary Figure S3** Distribution of COVID-19 case backfill across ultimate sizes. Illustrating the backfill patterns across different ultimate size categories: under 100, 100-1000, 1000-5000, 5000-10000, and over 10000. Four colors represent 75%, 85%, 95%, and 99% quantiles.

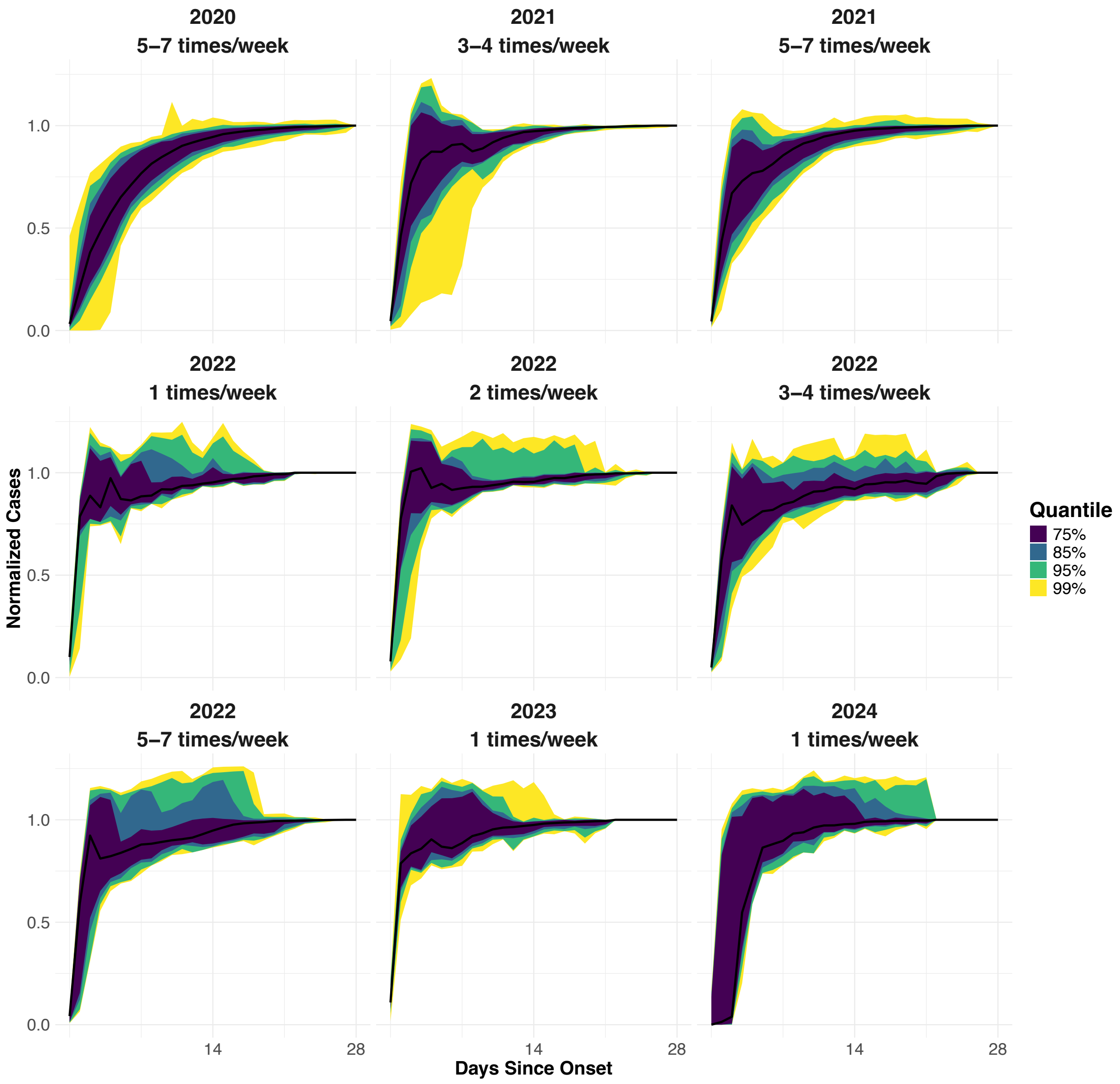


**Supplementary Figure S4** Distribution of COVID-19 case backfill under different reporting frequencies. Illustrating the backfill patterns across varying reporting frequencies observed in different years: 5-7 times, 3-4 times, 2 times, and 1 time per week. Four colors represent 75%, 85%, 95%, and 99% quantiles.

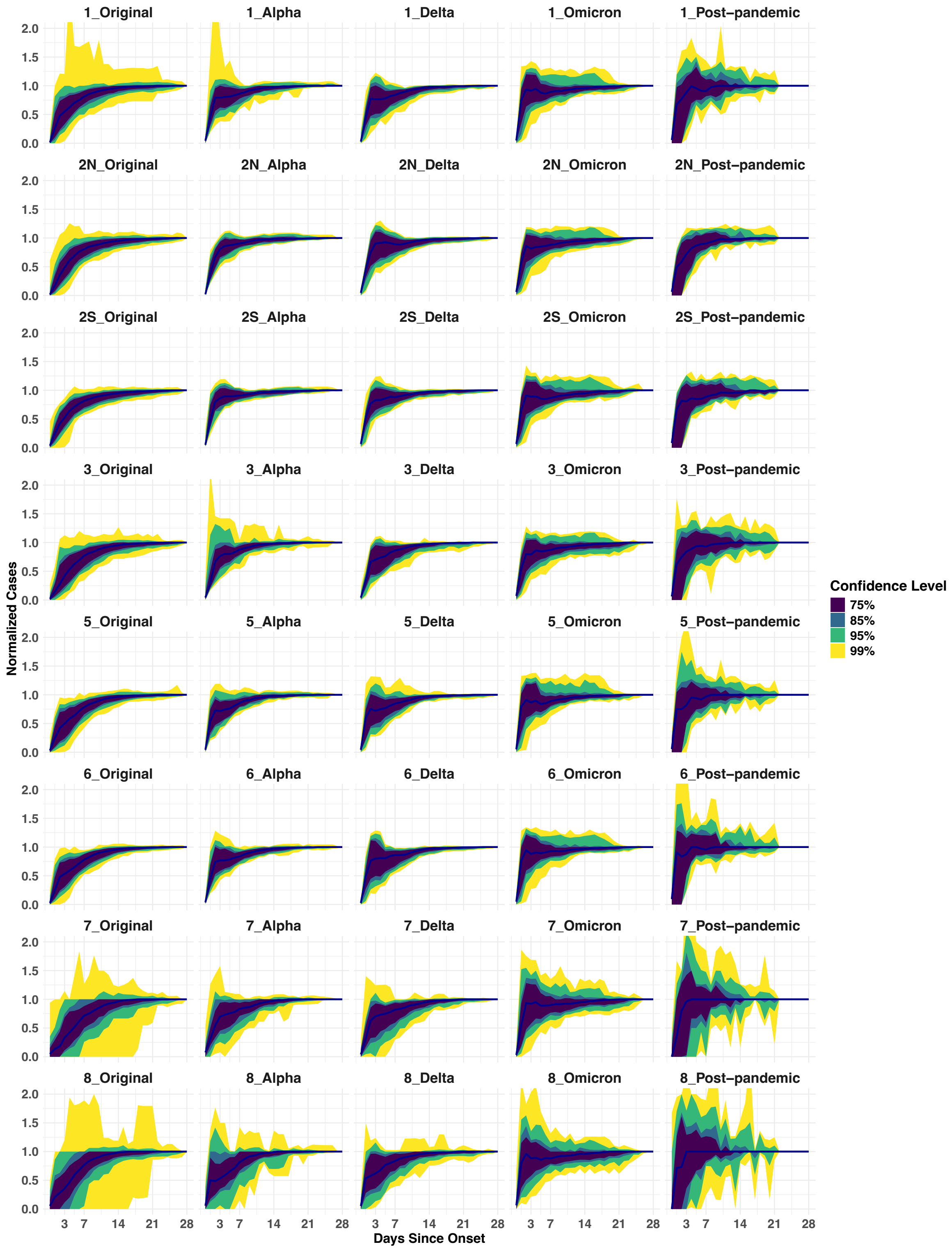


**Supplementary Figure S5** Regional distribution of COVID-19 case backfill across waves. Displaying the backfill patterns in the Original, Alpha, Delta, Omicron, and post-pandemic waves across eight emergency preparedness regions. Each row represents a specific region and each column corresponds to a wave. Four colors represent 75%, 85%, 95%, and 99% quantiles.

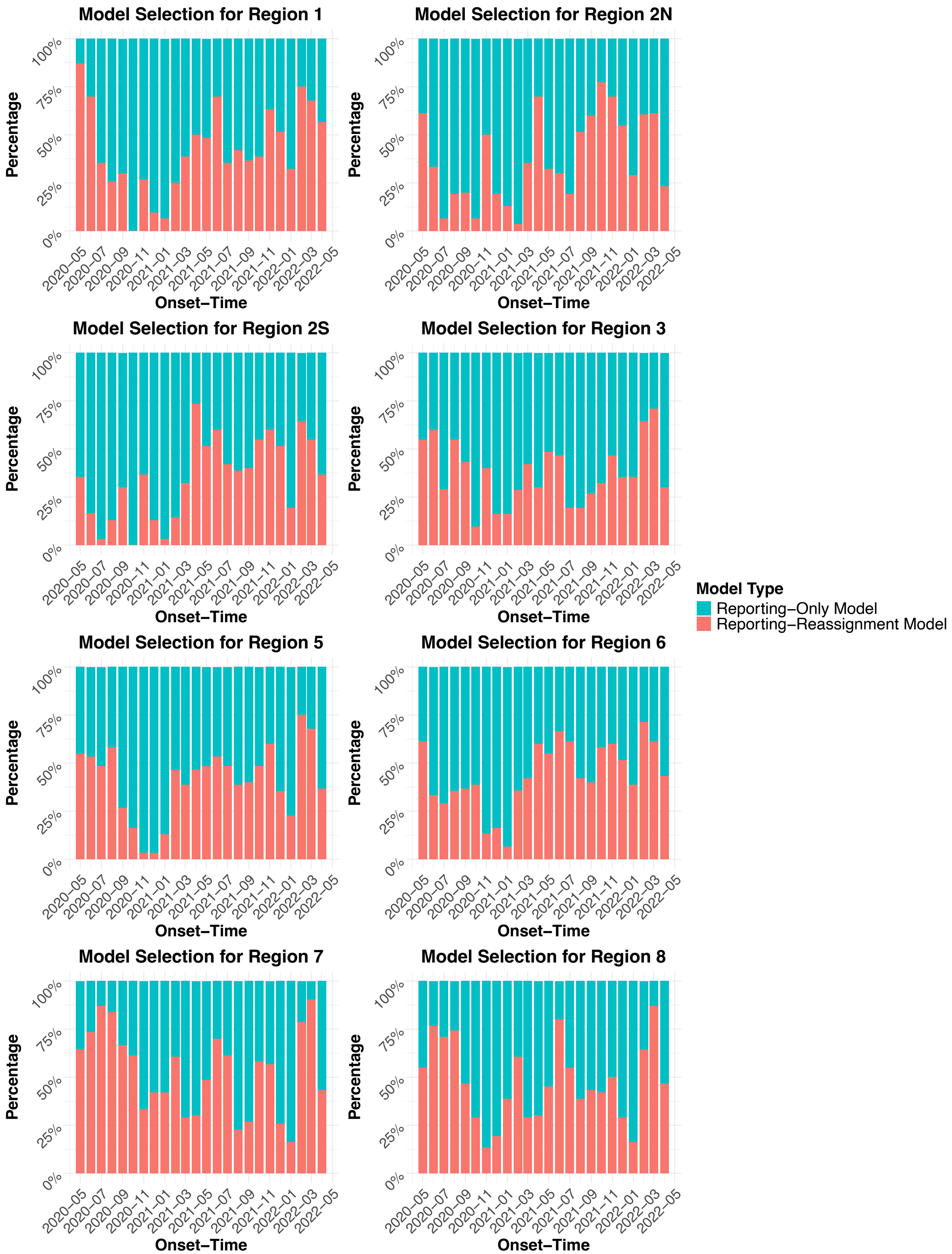


**Supplementary Figure S6** Model selection across regions. Showing monthly percentages of the basic and exponential-gamma models across eight emergency preparedness regions, with two colors representing the different models

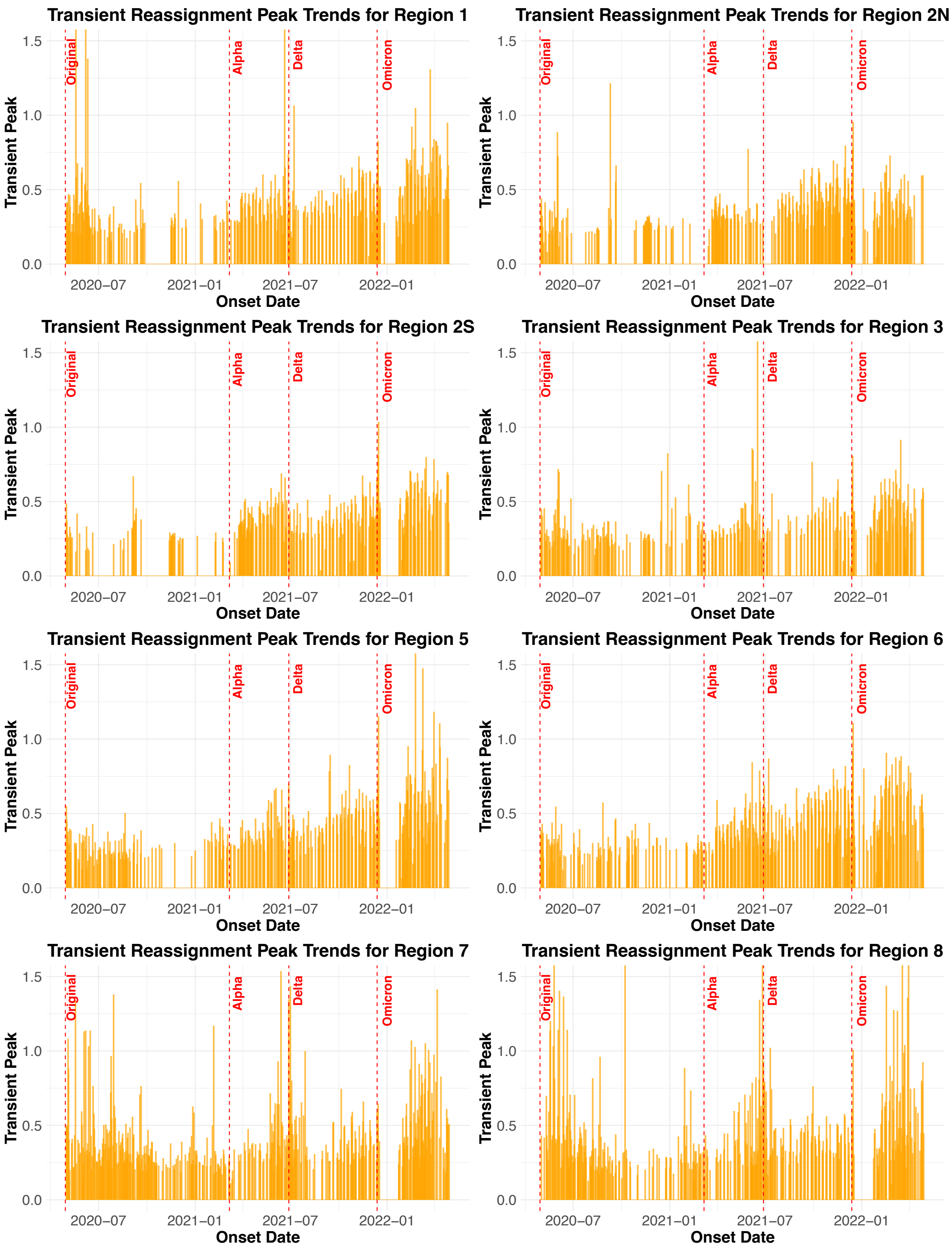


**Supplementary Figure S7** Transient reassignment peak trends across regions. Illustrating the relative transient peak in the reassignment process over time across eight emergency preparedness regions, with blanks representing dates selecting the basic model. Four vertical red dashed lines mark the start of a new wave in panels.

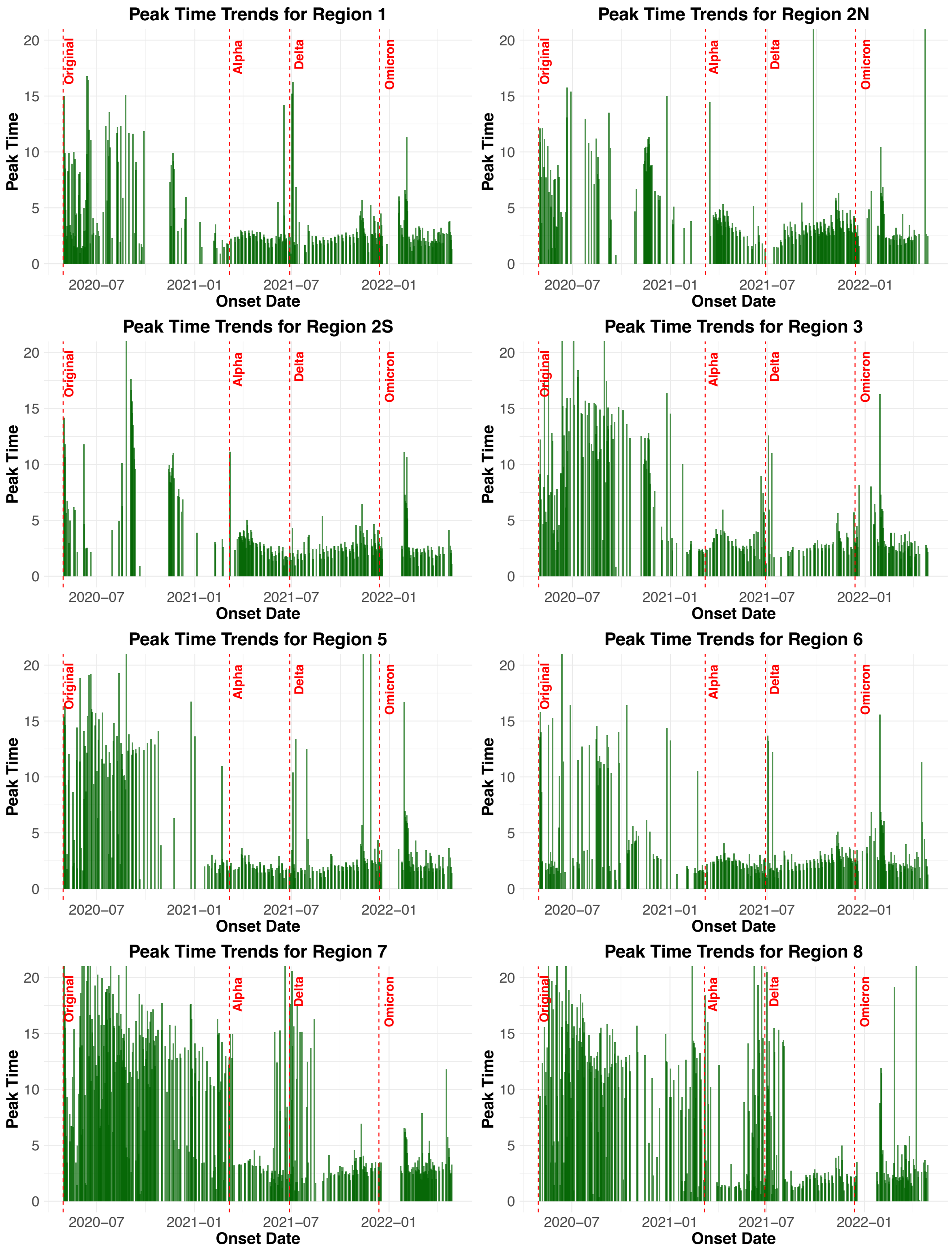


**Supplementary Figure S8** Peak time trends across regions. Illustrating trends of peak times in the reassignment process over time across eight emergency preparedness regions, with blanks representing dates selecting the basic model. Four vertical red dashed lines mark the start of a new wave in panels.

**Sensitivity analysis**

**(1) Methods**

Cases reported on the same day as their onset dates (Day 0) are impacted by many factors, such as reporting routines, work hours, and the timing of data extraction. As a result, Day 0 data may be less reliable than data from subsequent days. To examine if including Day 0 data affected parameter estimates, we conducted a sensitivity analysis where Day 0 data were excluded from the maximum likelihood estimation. This analysis allowed us to assess whether excluding Day 0 data could provide more reliable estimates or if including the initial data meaningfully improved parameter estimation.

**(2) Results**

The overall increase trends of reporting speeds were similar with or without Day 0 data. However, excluding Day 0 data affected model selection and parameter magnitudes differently across waves (Supplementary Figure S9). In the original wave, the reporting-only model remained dominant, and parameter estimates did not change substantially. In Alpha and Delta waves, the reporting-reassignment model was selected less frequently, and the exponential rate increased to 0.4. In the Omicron wave, although model selections were unchanged, the exponential rate was again elevated to 0.4. This higher rate suggested that 90% of cases could be captured within five days, which is inconsistent with real-world conditions. [1, 2] The sensitivity analysis indicates that excluding Day 0 data may reduce the model's ability to distinguish between delayed and reassigned cases, ultimately overestimating reporting speed.

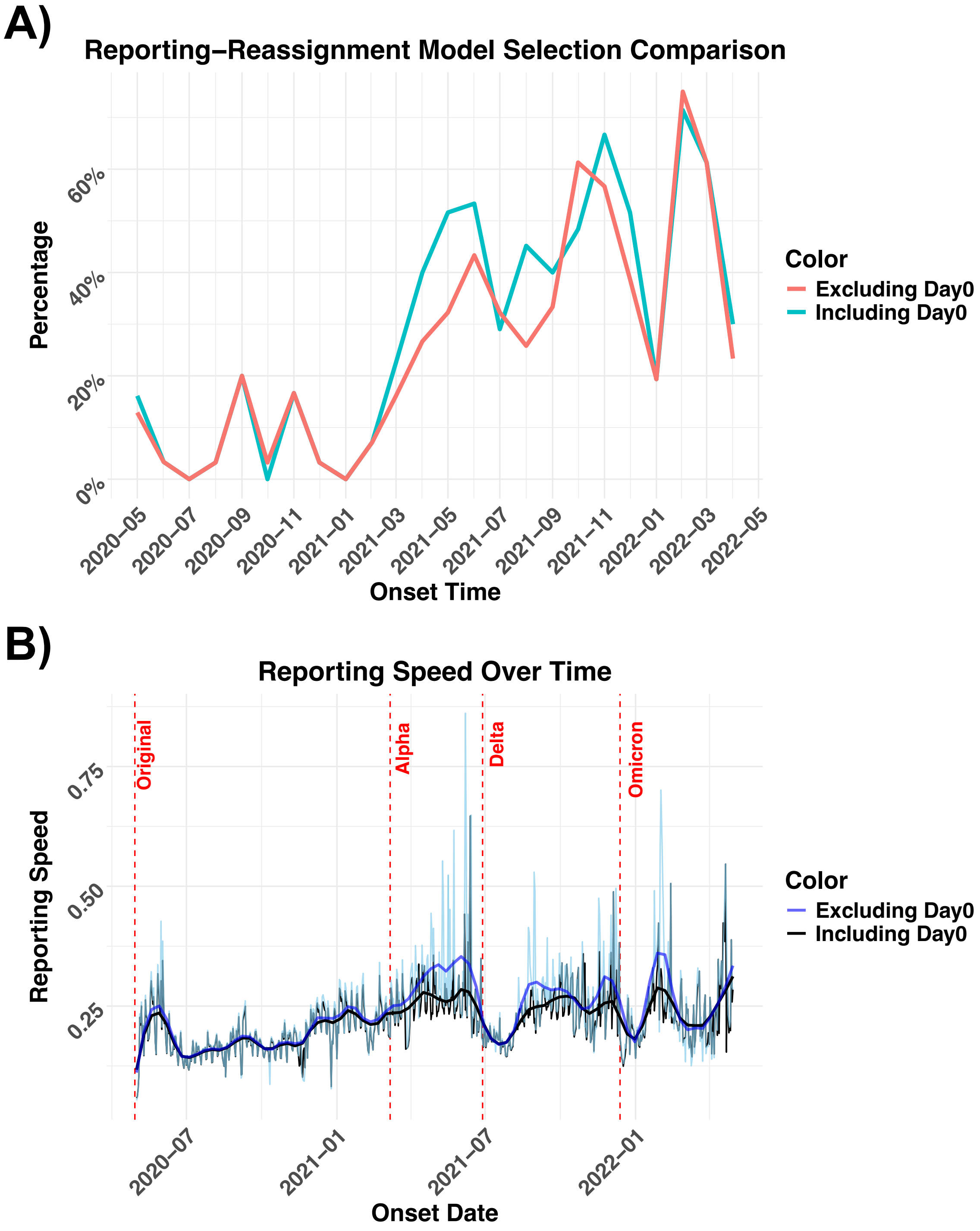


**Supplementary Figure S9** Model selection and exponential rate from sensitivity analysis. A) compares the monthly proportion of onset dates selecting the reporting–reassignment model when Day 0 data is included (blue) versus excluded (red). B) compares the exponential rate over time between including and excluding Day 0 data, with four vertical red dashed lines marking the start of a new wave.
